# Specific Effects of Integrase Inhibitors on Gut Microbiota in Men Who Have Sex with Men with and without HIV

**DOI:** 10.1101/2025.02.13.25321741

**Authors:** Marta Rosas Cancio-Suárez, Jorge Díaz-Álvarez, Claudio Díaz-García, Luis Miguel Nieto Salas, Matilde Sánchez-Conde, Elena Moreno, Alejandro G. García-Ruiz de Morales, Laura Marín-Pedraza, Clara Crespillo-Andújar, María Fons-Contreras, Raquel Ron, Ana del Amo-de Palacio, Marta González-Sanz, Santiago Moreno, Sergio Serrano-Villar

## Abstract

**Background:** HIV infection and antiretroviral therapy (ART) influence gut microbiota, affecting inflammation, immune function, and systemic health. However, isolating the effects of integrase strand transfer inhibitor (INSTI)-based ART on gut microbiota is complicated by confounding factors, including HIV status, immunosuppression, and sexual behavior.

**Methods:** This study examined three cohorts of men who have sex with men (MSM): 1) HIV-negative individuals using post-exposure prophylaxis (PEP) (n=22), 2) PWH with <350 CD4 cells/μL before and after ART (n=21 and n=13, respectively), 3) PWH on long-term INSTI-based ART with >500 CD4 cells/μL (n=17). Fecal microbiota was analyzed through 16S rRNA sequencing, with functional profiling using PICRUSt2. To compare differences in bacterial abundance and functions, we used ANCOM-BC2.

**Results:** PWH showed significantly lower alpha diversity than HIV-negative participants, especially those with marked immunosuppression. Short-term ART in PEP users showed no significant impact microbiota, while beta diversity clustered by HIV status rather than ART exposure. Pro-inflammatory taxa, such as *Prevotellaceae,* were enriched in PWH, reflecting interactions between HIV and MSM status. Functional profiling indicated elevated genes linked to antibiotic resistance, metabolism, and stress in PWH. While INSTI-based ART caused minor functional changes, it increased beneficial genera like *Barnesiella*.

**Conclusions:** While HIV significantly disrupts gut microbiota, INSTI-based ART preserves microbial diversity and community structure. Complementary microbiota-targeted interventions could enhance health outcomes for PWH.

## INTRODUCTION

People living with HIV (PWH) exhibit distinct gut microbiota profiles, which are associated with inflammation, transmission, and comorbidities (1–3). Initially, these changes were attributed to HIV’s direct impact on gut-associated lymphoid tissue (GALT), leading to a profound depletion of CD4 T cells that persists despite optimal antiretroviral therapy (ART)(4–6). This disruption causes bacterial translocation and chronic immune activation. However, current evidence highlights that other factors, such as sexual preference among men who have sex with men (MSM) (7–9) or geography, diet or the grade of immunosupression (10–12), also influence microbiota composition, perhaps more than HIV itself.

Methodological variability in microbiota studies adds complexity. Changes in bacterial composition in PWH—such as enrichment in *Enterobacteriaceae* and *Prevotella spp.* and reductions in *Bacteroides spp.*—are inconsistently reported, likely due to differences in sampling materials, analytical platforms, and study designs (13,14). Microbiota diversity is also controversial, as both increases, (15) and decreases (16) are reported in the literature.

More specifically, the role of ART in microbiota disruption remains unclear. While some studies show ART mitigates dysbiosis in PWH (7), others report minimal or even detrimental effects (17–19). INSTI-based ART has been associated with lower systemic inflammation, higher alpha diversity, and distinct bacterial profiles compared to other regimens (20), suggesting potential benefits in reducing chronic immune activation and promoting gut microbial resilience in the context of HIV treatment. However, isolating its effects remains challenging due to ART combinations, frequent treatment modifications, and inherent difficulties in controlling prescription bias, as patients receiving protease inhibitors often differ in lifestyle factors such as dietary habits, exercise routines, and substance use, all of which influence the microbiota, from those initiating INSTI-based regimens.

Understanding how ART affects the gut microbiota is necessary to assess its role in HIV progression, comorbidities, and chronic inflammation. In this study, we aim to control the specific impact of INSTI-based ART on the gut microbiota composition and function under varying conditions of HIV status and immune recovery by controlling for the confounding effects of sexual practices, type of ART, and immunosuppression.

## MATERIAL AND METHODS

We conducted a single-center observational study to investigate how integrase inhibitors influence the gut microbiome in healthy MSM receiving INSTI-based post-exposure prophylaxis (PEP). Furthermore, we aimed to compare the differences in the intestinal microbiota between healthy MSM, HIV+ MSM on ART with long-term viral suppression and >500 CD4 cells/μL and PWH not receiving ART and CD4 cell count <350/μL or Acquired Human Immunodeficiency Syndrome (AIDS).

### Sample and cohort collection

This study included three cohorts (**figure 1**). **All participants were MSM:**

1. PEP (HIV- PrePEP and HIV- PostPEP): 22 healthy MSM from the Sexual Health Clinic undergoing PEP. Two stool samples were obtained per participant: one before starting PEP and one after 28 days (treatment completion).
2. Advanced HIV (HIV+ lowCD4 preART and HIV+ lowCD4 postART): 23 MSM with advanced HIV infection (AIDS or CD4 <350 cells/uL at diagnosis) providing samples before starting ART and 48 weeks after starting ART.
3. Stable ART (HIV+ highCD4 postART): 17 asymptomatic MSM with HIV on stable INSTI-based ART for an average of 9.5 years.

**Figure 1:**
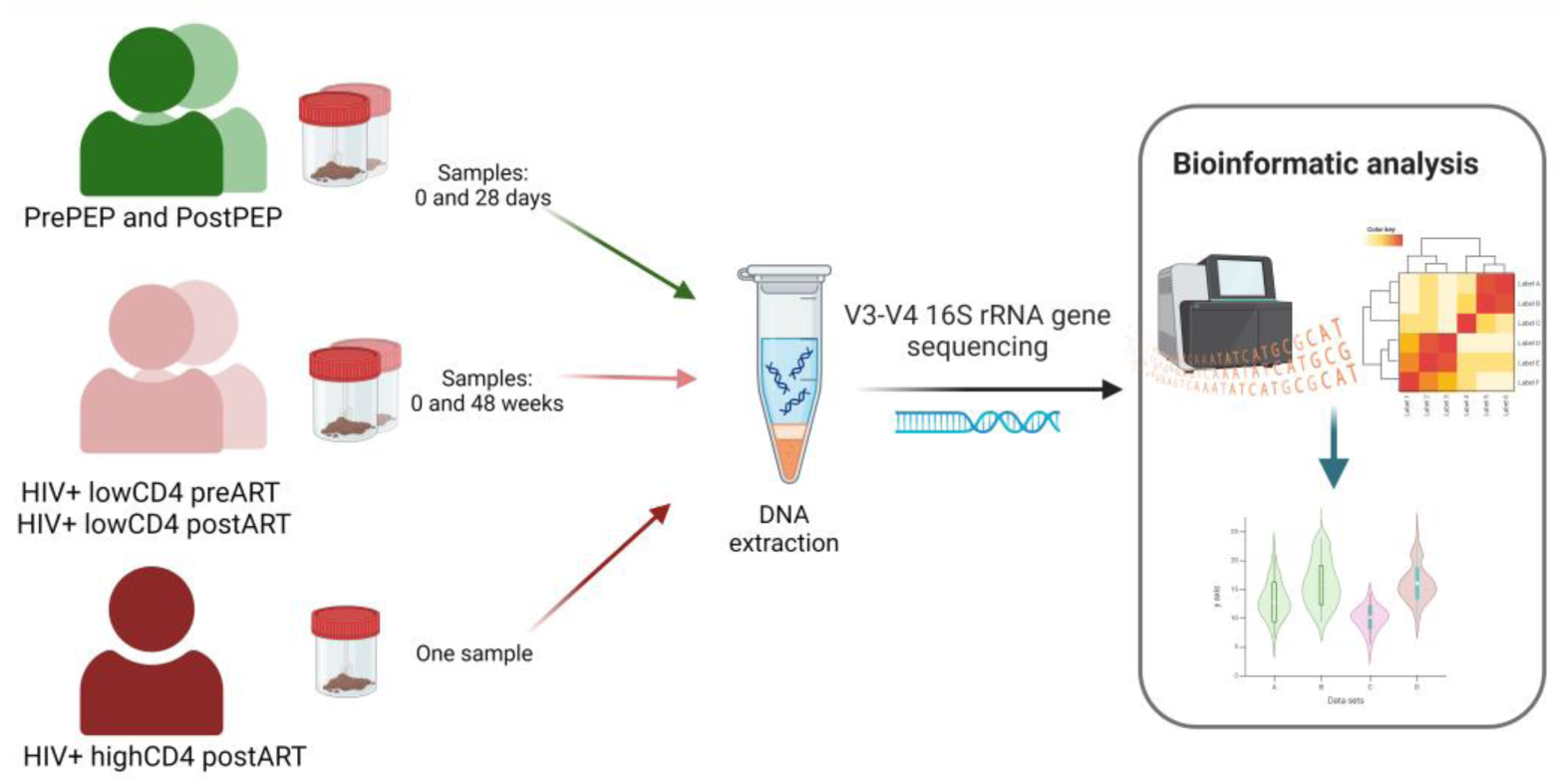
Representation of study design. All participants were MSM with different HIV, ART or CD4 counts status.

Exclusion criteria included age under 18, prior inflammatory bowel disease, autoimmune disease, cirrhosis, or ART exposure in the PEP cohort.

### Bioinformatics analysis

Fecal samples were stored in RNAlater (Life Technologies) at −80°C until processed. DNA was extracted using PowerFecal Pro DNA Kit (Qiagen) and quantified by Qubit fluorometry. The V3-V4 region of the 16S rRNA gene was amplified and sequenced using the Illumina MiSeq platform.

We processed the data using ampliseq pipeline (v2.9.0) (21). Raw sequencing reads were subjected to quality filtering to remove unwanted sections of the sequences. Valid sequences were analyzed using the DADA2 algorithm (22), identifying 5917 unique amplicon sequence variants (ASVs). Taxonomic assignment was conducted using SILVA database (v138) (23) with Naive Bayes classifier produced by QIIME2 2023.7. We used ANCOM-BC2 for differential abundance analysis (24). Absolute abundance tables were aggregated to the Genus level and used as input. Differences were expressed in log fold change and p-values were corrected for multiple testing by false discovery rate.

For the predicted functional analysis of bacterial communities, we used Phylogenetic Investigation of Communities by Reconstruction of Unobserved States 2 (PICRUSt2) (25). This tool uses genomic databases and the phylogeny of known organisms to predict the presence of gene functions in an environmental sample. We then analyzed the predicted absolute abundances of Kyoto Encyclopedia of Genes and Genomes (KEGG) orthologs (KO) and enzyme classification (EC) numbers with ANCOM-BC2. We applied a minimum threshold of 2 log fold changes to consider a feature as functionally relevant. We corrected p-values by a false discovery rate. We used R version 4.2 and RStudio version 2024.04.2 (26) for statistical analysis and graphics production.

### Statistical analysis

The study’s design allowed us to explore how different factors influence the gut microbiota through targeted comparisons. To assess the effect of HIV infection, we compared the microbiota of HIV- before PEP initiation with that of HIV+ with advanced disease (CD4 <350) before ART. To evaluate the impact of immunosuppression, we examined differences between HIV+ with advanced disease (CD4 <350) after ART and those with stable immune recovery (CD4 >500) on long-term ART. Finally, to isolate the effects of ART itself, we analyzed microbiota changes in HIV-negative MSM before and after completing PEP.

We used Chi-square tests for categorical variables, and Mann-Whitney for continuous variables. For alpha and beta diversity analyses, we applied Kruskal-Wallis, Wilcoxon rank sum, and PERMANOVA tests.

The sample size recommendations for microbiota studies suggest that at least 20 subjects per group be included to detect differences when present (27)

### Ethics

The ethics committee of the Ramón y Cajal University Hospital (, Protocol Number 052/18) approved the study. Before the study procedures, all participants signed informed consent. The protocol was registered with ClinicalTrials.gov with the Identifier NCT04460924.

## RESULTS

We included 62 MSM: 22 Pre and PostPEP, 23 HIV+ lowCD4 preART and postART, and 17 HIV+ highCD4 postART. Table 1 summarizes their baseline characteristics. The 62 study subjects provided 95 samples, which were subsequently processed for the analysis of microbiota composition.

**Table 1.** Population baseline characteristics by group.

|  |  | HIV+ lowCD4 | HIV+ highCD4 postART | PEP | p-value |
| --- | --- | --- | --- | --- | --- |
| **N** |  | 23 | 17 | 22 |  |
| **Age, median (IQR)** |  | 36.5 (30.5-43.1) | 42.7 (36-53.3) | 37 (31-44) | 0.16 |
| **Body mass index (kg/m2), median (IQR)** |  | 23.4 (21.9-26.1) | 24.6 (23.5-26.9) | 24.4 (22.2-25.7) | 0.31 |
| **HCV status at diagnosis, n (%)** | Negative | 20 (87%) | 16 (94%) | 22 (100%) | 0.20 |
|  | Positive | 3 (13%) | 1 (6%) | 0 (0%) |  |
| **AIDS at diagnosis, n (%)** | Yes | 2 (9%) | 2 (12%) | 0 (0%) | <0.001 |
| **Nadir CD4, median (IQR)** |  | 267 (138-340) | 249 (113-300) | N/A | 0.55 |
| **Current CD4, median (IQR)** |  | 267 (138-340) | 609 (465-704) | N/A | <0.001 |
| **PreART CD8, median (IQR)** |  | 1117 (711-1525) | 276 (210-392) | N/A | <0.001 |
| **Current CD4/CD8 count** |  | 0.22 (0.13-0.37) | 0.73 (0.53-0.85) | N/A | <0.001 |
| **Viral load at diagnosis (copies/ml), median (IQR)** |  | 63000 (27839-155131) | 92320 (9210-422000) | N/A | 0.42 |
| **Antibiotics in the last 3 months** | No | 17 (74%) | 14 (82%) | 22 (100%) | 0.042 |
|  | Yes | 6 (26%) | 3 (18%) | 0 (0%) |  |

Abbreviations: MSM, men who have sex with men; IQR, interquartile range; HCV, Hepatitis C virus; AIDS, acquired immune deficiency syndrome; ART, antiretroviral treatment; ART, antiretroviral treatment; N/A, not applicable

### Differences in alpha and beta diversity across groups

Alpha diversity indices, which measure the variety of species within a sample, varied significantly between groups (**Figure 2**). The HIV- group displayed the lowest diversity compared to HIV+ groups, irrespective of ART status. Principal Coordinate Analysis (PCoA) of weighted UniFrac distances, which quantifies compositional differences between samples based on the presence and abundance of microbial taxa, revealed significant differences in microbiota composition among groups (global PERMANOVA, p < 0.001) (**Figure 3**). HIV- participants formed a distinct cluster, separated from HIV+ >500 CD4 individuals (p < 0.001). Importantly, there was no effect of one month of INSTI-based ART (PEP group) on alpha diversity.

**Figure 2:**
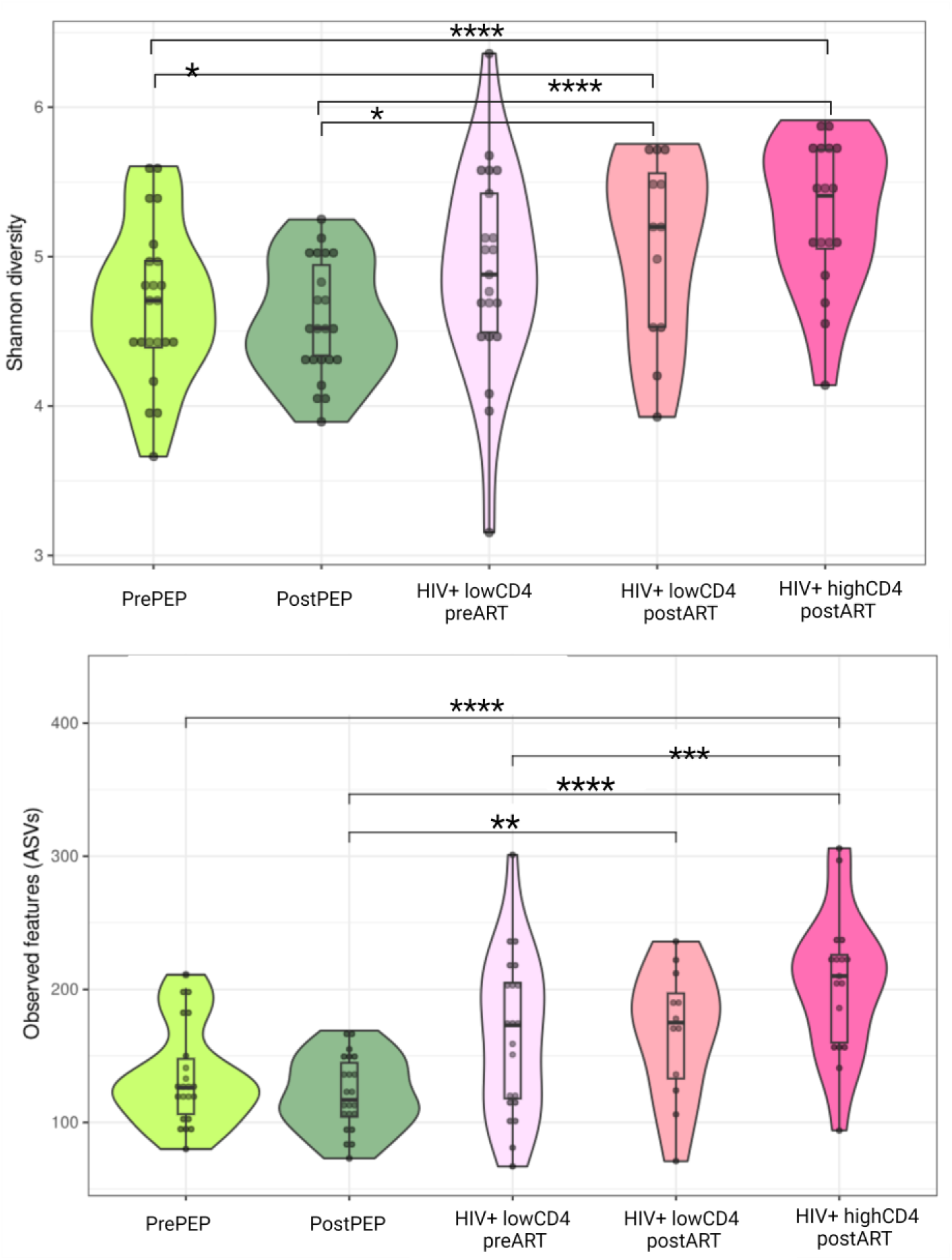
Violin plots depicting Shannon entropy (upper panel) and Observed features (lower panel) indexes representing the alpha diversity across five groups of MSM: PrePEP, PostPEP, HIV+ lowCD4 preART, HIV+ lowCD4 postART, and HIV+ highCD4 postART. Subgroups of MSM without HIV starting PEP are depicted in green tonalities, while MSM with HIV and different ART and CD4 status are represented in pink tonalities. Statistical comparisons (Wilcoxon test) are shown with significance levels indicated by an asterisk. Differences not marked with a star were not statistically significant. The asterisks represent the following significance levels: * p < 0.05, ** p < 0.01, *** p < 0.001, **** p < 0.0001.

**Figure 3:**
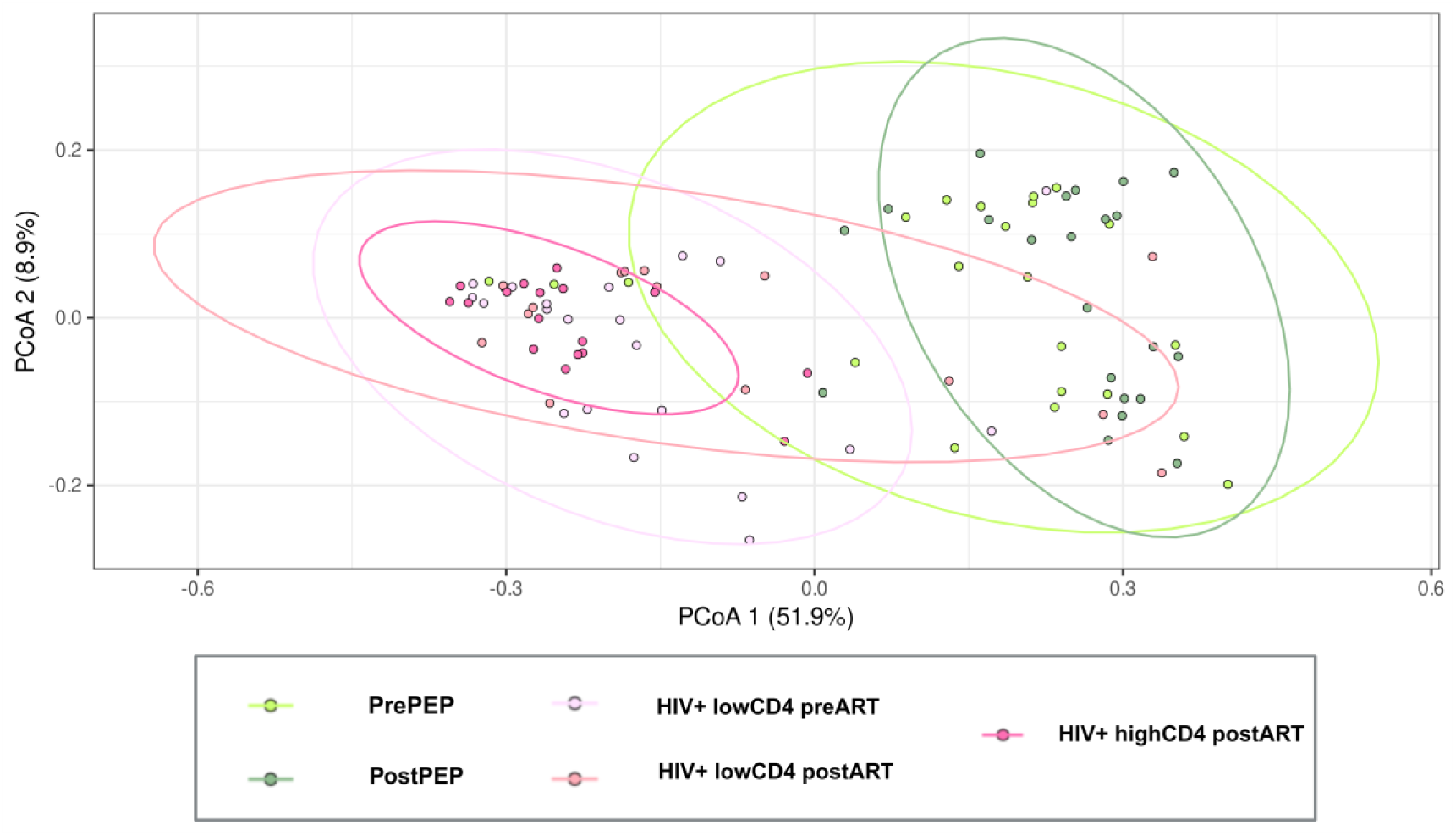
PCoA based on the weighted Unifrac distance of the different groups of MSM. Note: Those who are PWH are differentiated in shades of pink, and those who are not shades of green.

### Specific microbial taxa differed between study groups

We observed distinct patterns in microbiota composition when comparing specific groups (**Figure 4** and **Table S1**).

**Figure 4:**
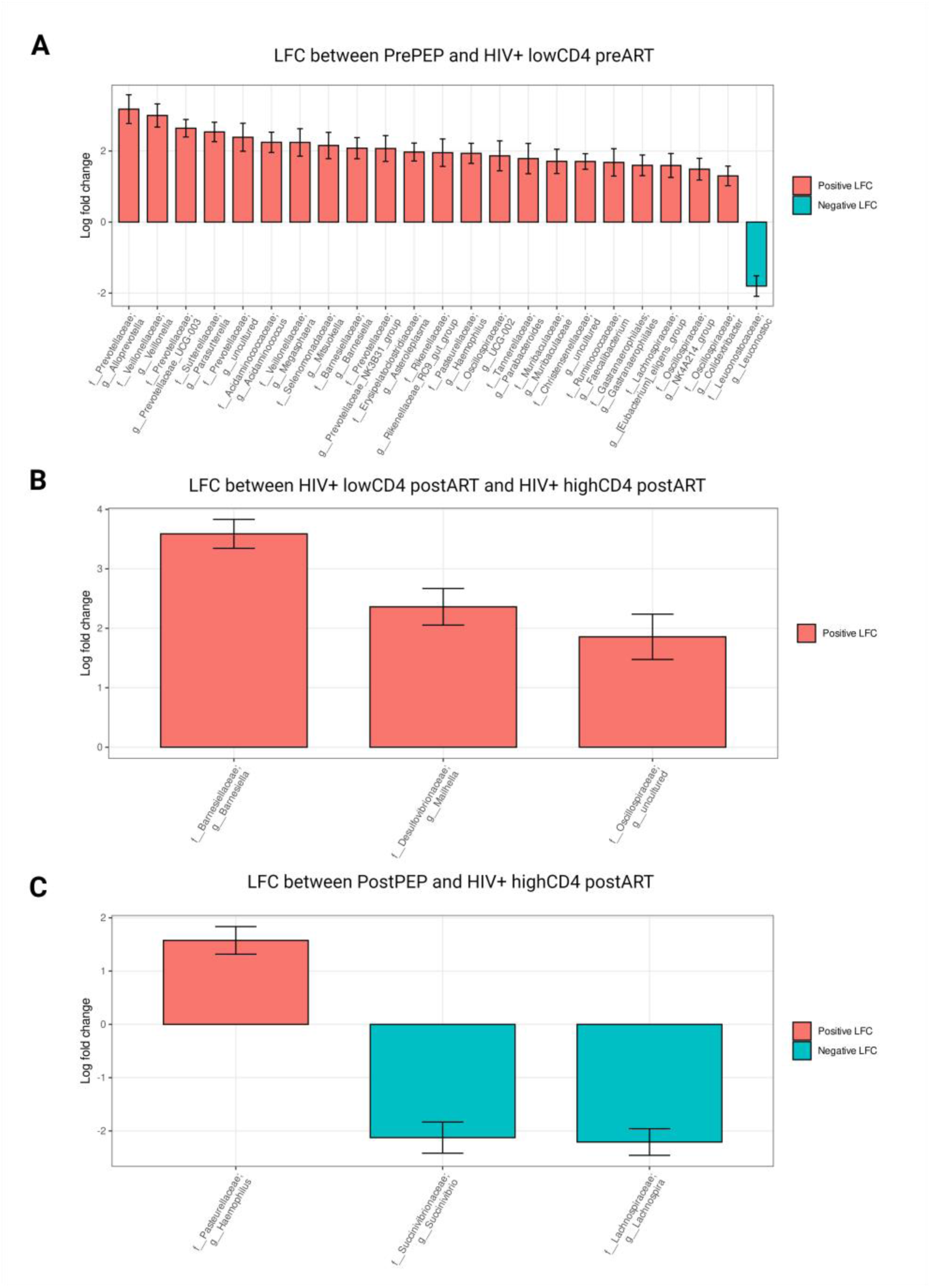
Log fold changes (LFC) in gut microbial taxa when comparing different groups of MSM. A.LFC between HIV-prePEP and HIV+ lowCD4 preART. B LFC between HIV+ lowCD4 postART and HIV+ highCD4 postART. C LFC between HIV- prePEP and HIV- postPEP. Note: The bar graph illustrates the mean LFC (± standard error) of relative abundances for the differential taxa following ANCOMB-BC2 analysis. Positive LFC values indicate taxa enriched in individuals undergoing long-term ART compared to short-term ART. Error bars represent the standard error of the mean.

First, in the comparison between HIV- individuals before PEP initiation and PWH individuals with advanced disease (<350 CD4), we noted a marked enrichment of several bacterial families in the latter, including *Prevotellaceae*, *Veillonellaceae*, *Sutterellaceae*, *Acidaminococcaceae*, and *Selenomonadaceae*. Genera such as *Alloprevotella* and unclassified *Prevotellaceae* stood out as significantly more abundant, whereas *Leuconostoc*and unclassified *Peptostreptococcaceae* genera decreased. In addition to those in **Table 2**, another twelve families had greater abundance in PWH without ART with minor differences (log fold changes — LFC— < 1.5)(**Figure 4A**).

Next, among MSM with HIV on ART, the microbiota of those with CD4 counts >500 showed slight increases in genera such as *Barnesiella* and *Mailhella* (**Figure 4B**). These differences, although minor, suggest some degree of microbial modulation associated with better immune recovery.

Finally, when comparing microbiota within the MSM HIV- cohort following PEP, only one genus, *Haemophilus*, increased, while *Lachnospira* and *Succinivibrio* abundances decreased (**Figure 4C**).

**Figure S1** shows the relative abundance and prevalence of the differential bacterial taxa across groups.

In the context of the previous results showing a drastic impact of HIV-induced immunosuppression on gut microbiota structure, this analysis highlights the relatively minor effects of INSTI-based ART on gut microbiota composition, underscoring the contrast between the two factors.

### Functional roles of microbial taxa across groups

We sought to investigate the functional roles of the bacterial taxa identified, using PICRUSt2 to predict the abundance of KEGG orthologs and enzymes and ANCOM-BC2 to detect significant differences across groups (Figure 5). Our findings revealed the following patterns.

**Figure 5:**
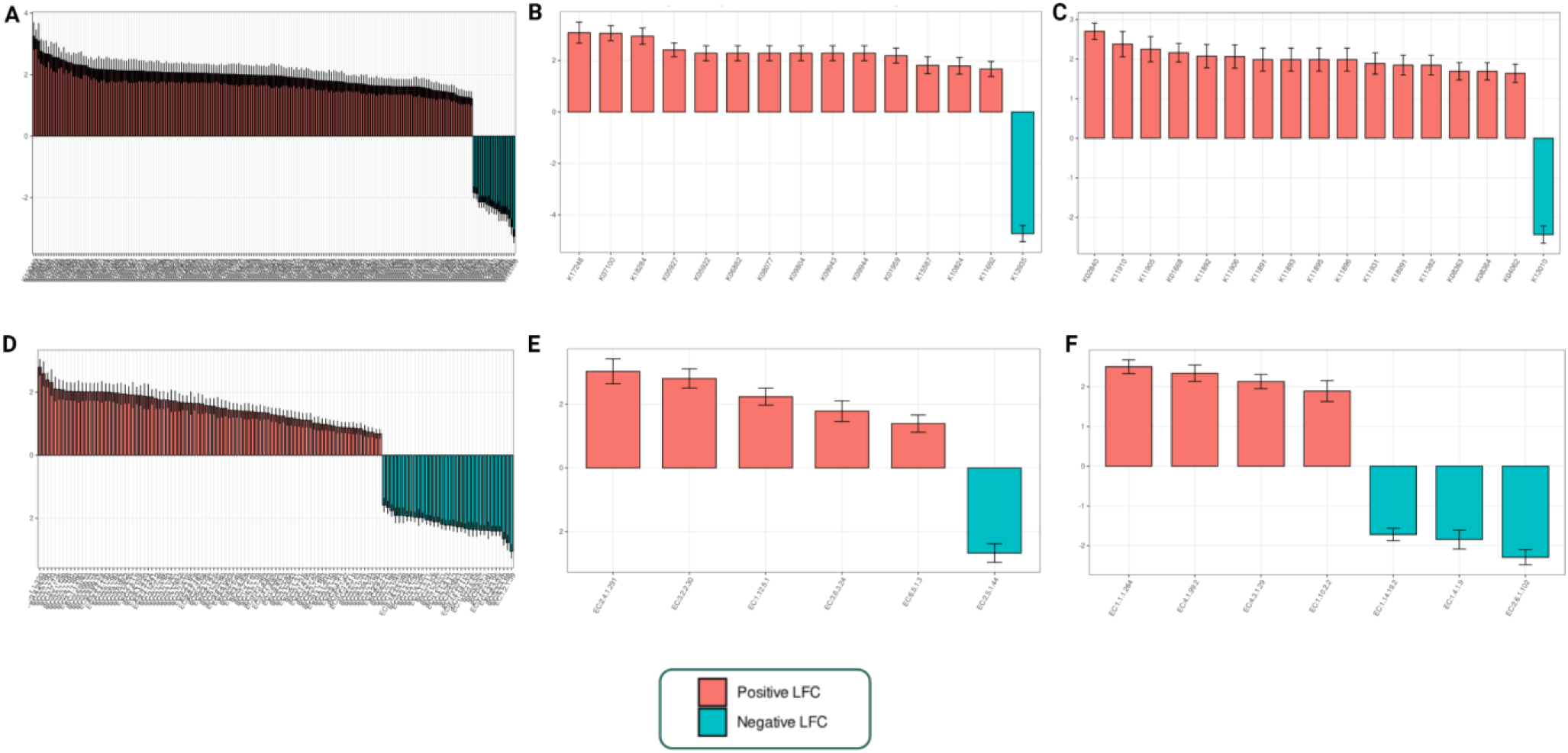
Functional terms differentially found in the three cohorts of MSM. Upper images: KEGG were obtained with Picrust2, compared between groups with ANCOM-BC2, and selected based on their log-fold change (LFC). **A.** Comparison of KO terms between PrePEP and HIV+ lowCD4 preART. **B.** Comparison of KO terms between HIV+ lowCD4 postART and HIV+ highCD4 postART. **C.** Comparison of KO terms between PrePEP vs. PostPEP. **Lower images:** EC terms obtained with Picrust2, compared between groups with ANCOM-BC2 and selected based on their LFC. **D.** Comparison of EC terms between PrePEP and HIV+ lowCD4 preART. **E.** Comparison of EC terms between HIV+ lowCD4 postART and HIV+ highCD4 postART. **F.** Comparison of EC terms between PrePEP vs. PostPEP. **Note:** Genus in red have a positive LFC (higher abundance) in the reference group. Genus in blue have a negative LFC (lower abundance) in the reference group.

When comparing HIV- individuals before ART and PWH with advanced disease (<350 CD4), we observed a striking divergence, with over 193 KEGG Orthologs (KO) and 123 Enzyme Classification (EC) terms showing significant differences. Untreated PWH displayed higher abundance in bacterial genes associated with cell wall synthesis (*murU*), energy metabolism (*citT*), and environmental response mechanisms (*TtrS*). Conversely, genes involved in bacterial conjugation (*trbC*) were less abundant. Significant increases in enzymes such as benzil reductase and endopeptidase, linked to aromatic compound metabolism, were also noted, alongside reductions in isomerases, decarboxylases, and dehydrogenases (**tables S2** and **S3**).

Comparing HIV+ individuals with advanced disease (<350 CD4) to those with higher immune recovery (>500 CD4) after ART, we identified 15 KO terms and 6 EC terms with significant differences. Individuals with better immune recovery showed enrichment in genes involved in cell surface modification (*pglJ*) and nucleotide synthesis (*K07100*), while genes related to fatty acid metabolism (*mdcH*) were less abundant. Enzymes supporting redox processes and detoxification (4-alpha-N- acetylgalactosaminyltransferas and aminodeoxyfutalosine nucleosidase) were more prevalent in individuals with higher CD4 counts (**tables S2** and **S3**).

For HIV- individuals, pre- and post-ART comparisons revealed fewer differences, with 17 KO terms and 7 EC terms showing notable shifts, suggesting that INSTI-based ART has a relatively minor impact on gut microbiota functions. After ART, genes linked to cell envelope biosynthesis (*waaB*, *rfaB*) and carbohydrate metabolism increased, while those associated with virulence (*per*, *rfbE*) decreased. Additionally, enzymes related to detoxification and stress response became more abundant post-ART. These findings suggest that INSTI-based ART exerts a minor yet beneficial impact on gut microbiota functions, highlighting its potential for preserving microbial balance during treatment.

## DISCUSSION

Our study highlights key findings about the gut microbiota in MSM HIV- and MSM living with HIV on INSTI-based ART. Specifically, we found that i) alpha diversity differed according to HIV and ART status, but short-term ART had no significant effect on alpha diversity; ii) beta diversity analyses showed clustering according to HIV status, reinforcing the idea that HIV, rather than ART, drives the differences in gut microbiota composition; and iii) functional analyses revealed distinct patterns of microbial gene abundance in PWH, emphasizing alterations linked to infection, immune recovery, and a minor impact of ART on microbiota functions.

Alpha diversity analyses revealed differences in PWH compared to healthy controls, with the most pronounced differences observed in immunosuppressed individuals. Notably, no significant changes were observed within the HIV- group (PEP users) after one month of INSTI-based ART, supporting the idea that INSTI-based ART has minimal ecological impact (28–30). Beta diversity further supported these findings, showing apparent clustering based on HIV status rather than ART exposure (30,31). These results emphasize the role of HIV in shaping gut microbiota, independently of MSM status, and reinforce the limited impact of short-term ART.

Understanding how ART affects gut bacteria is challenging because ART improves immune function, and there are many different treatment combinations and selection factors to consider. Available research findings are contradictory. In keeping with our, some long-term studies found minimal changes in gut bacteria when patients started ART, whether they used INSTI or non-nucleoside reverse transcriptase inhibitors (32). This was true even for patients with advanced HIV disease who were followed for 48 weeks (33). However, location and initial gut bacteria composition seem to matter: a study from Zimbabwe found that in rural areas, combining ART with cotrimoxazole reduced bacterial diversity after 6 months, especially in people who started with more diverse gut bacteria (34). Beyond the impact of ART on immune reconstitution and on gut microbiota, drugs themselves might directly affect bacterial diversity. Supporting this idea, laboratory research has revealed that some antiretroviral drugs can actually kill bacteria, including hard-to-treat resistant bacteria found in the gut and the vagina (35).

Our findings align with research showing enrichment of *Prevotellaceae* in PWH, a family linked to pro-inflammatory processes and associated with various inflammatory conditions. In MSM, *Prevotellaceae* enrichment is a well-documented trait, and its pronounced presence in PWH may reflect a combined effect of HIV-associated immune dysregulation and MSM-related microbiota patterns, contributing to dysbiosis and chronic inflammation (27,30). This pro-inflammatory family is often associated with MSM but in our cohort it was more abundant in PWH, suggesting a potential synergistic effect between HIV and MSM status on *Prevotellaceae* abundance (36). Other inflammatory-associated families, such as *Veillonellaceae* and *Sutterellaceae*, were also elevated in PWH, reinforcing the link between HIV and microbiota dysbiosis (37–39). We have previously commented on the lack of significant changes in bacterial taxa before and after short-term ART in PEP users. However, the observed increase in beneficial genera like *Barnesiella* in patients with higher CD4 counts suggests a potential restorative effect of INSTI-based ART on the microbiota of PWH (31).

Understanding the impact of ART on the microbiota may help clarify the mechanisms by which HIV infection and its treatment influence systemic inflammation, immune recovery, and overall health outcomes. Functional predictions revealed significant changes in bacterial genes and pathways, with PWH showing increased genes related to antibiotic resistance (*murU*), altered metabolism (*citT*), and stress responses (*ttrS*) (40–44). These functional changes could influence clinical outcomes by enhancing bacterial resilience in a disrupted gut environment, exacerbating chronic inflammation, and altering metabolic interactions, potentially accelerating disease progression and complicating immune recovery in PWH. These changes may contribute to broader health implications by potentially enhancing bacterial survival in a disrupted gut environment, increasing inflammation, and modifying the metabolic interactions between the microbiota and host, all of which could exacerbate HIV-associated dysbiosis (46,46). Interestingly, HIV- individuals post-ART demonstrated an increase in genes linked to cell envelope biosynthesis (*waaB*, *rfaB*) and a decrease in virulence-associated genes, suggesting ART may modulate pathogenic traits within the microbiota (46,47).

The primary strength of our study lies in the homogeneous cohort, with all participants being MSM from the same urban area, minimizing confounding factors such as sexual behavior and gender. This uniformity allowed us to isolate the effects of HIV and ART more effectively, providing a clearer understanding of their specific impacts on gut microbiota composition and function. Controlling these variables is particularly important in microbiota studies, as both sexual behavior and gender significantly influence gut microbiota composition, potentially masking the specific effects of HIV and ART (16,48). Integrating taxonomic and functional analyses provided a more comprehensive view of microbiota changes (49). However, detailed dietary information was absent, and the limited sample size warrants a cautious interpretation of our results. Future longitudinal studies with larger sample sizes are needed to validate our findings and explore the long-term effects of INSTI-based ART on gut microbiota. Specifically, these studies should investigate the sustained impact on bacterial diversity, functional capacities such as metabolic and immune-related pathways, and the potential modulation of inflammation and gut barrier integrity in PWH.

In conclusion, our study provides new insights into the gut microbiota in MSM and PWH, demonstrating that while HIV infection significantly impacts microbiota diversity and function, short-term INSTI-based ART has a limited effect. The observed shifts in bacterial taxa and functional pathways highlight the complex interplay between HIV, immune status, and ART. Understanding these dynamics is necessary for developing microbiota-targeted therapies to improve health outcomes in PWH.

## Data Availability

All data produced in the present study are available upon reasonable request to the authors

## Acknowledgments

The authors thank all the study participants and their families and the staff involved in this study for their commitment to clinical research.

## Author’s contributions

All the authors reviewed and approved the manuscript.

## Data availability

The raw reads from 16S rRNA gene sequencing are publicly available in the European Nucleotide Archive (ENA) under the project accession PRJEB84358. Authors declare that all other data supporting the study’s findings are available from the corresponding authors on reasonable request.

