## supplemental files for "Specific Effects of Integrase Inhibitors on Gut Microbiota in Men Who Have Sex with Men with and without HIV"

Supplementary Material Index

### ***Table S1: LFC of differential abundances between prePEP  vs HIV+ lowCD4 preART***

| Taxon (families and genera) | LFC | SE | P-value | Adjusted P |
| --- | --- | --- | --- | --- |
| Prevotellaceae; gender *Alloprevotella* | 3.18 | 0.40 | 2.02E-08 | 3.52E-06 |
| Prevotellaceae; unidentified gender | 2.65 | 0.24 | 3.39E-05 | 5.62E-03 |
| Veillonellaceae; genre *Veillonella* | 2.60 | 0.31 | 7.55E-06 | 1.28E-03 |
| Sutterellaceae; genre *parachute* | 2.48 | 0.27 | 2.50E-04 | 3.80E-02 |
| Acidaminococcaceae; gender *Acidaminococcus* | 2.26 | 0.28 | 3.70E-06 | 6.36E-04 |
| Veillonellaceae; gender *Megasphaera* | 2.21 | 0.39 | 9.80E-05 | 1.56E-02 |
| Selenomonadaceae; gender *With Mitsuoke* | 2.17 | 0.37 | 1.47E-04 | 2.27E-02 |
| Prevotellaceae; unidentified gender | 2.16 | 0.35 | 1.56E-06 | 2.70E-04 |
| Prevotellaceae; unidentified gender | 2.08 | 0.25 | 7.31E-05 | 1.18E-02 |
| Prevotellaceae; unidentified gender | 2.05 | 0.36 | 1.29E-05 | 2.17E-03 |
| Leuconostocaceae; gender *Leuconostoc* | -1.85 | 0.28 | 1.37E-05 | 2.30E-03 |
| Peptostreptococcaceae; unidentified gender | -2.31 | 0.22 | 1.42E-04 | 2.20E-02 |

LFC: *Log fold change*; SE: Standard error; Corrected P: FDR-corrected P value.

Note: P and corrected P values are represented in scientific notation, which expresses numbers using a power of 10 and a coefficient (coefficient × 10^exponent).

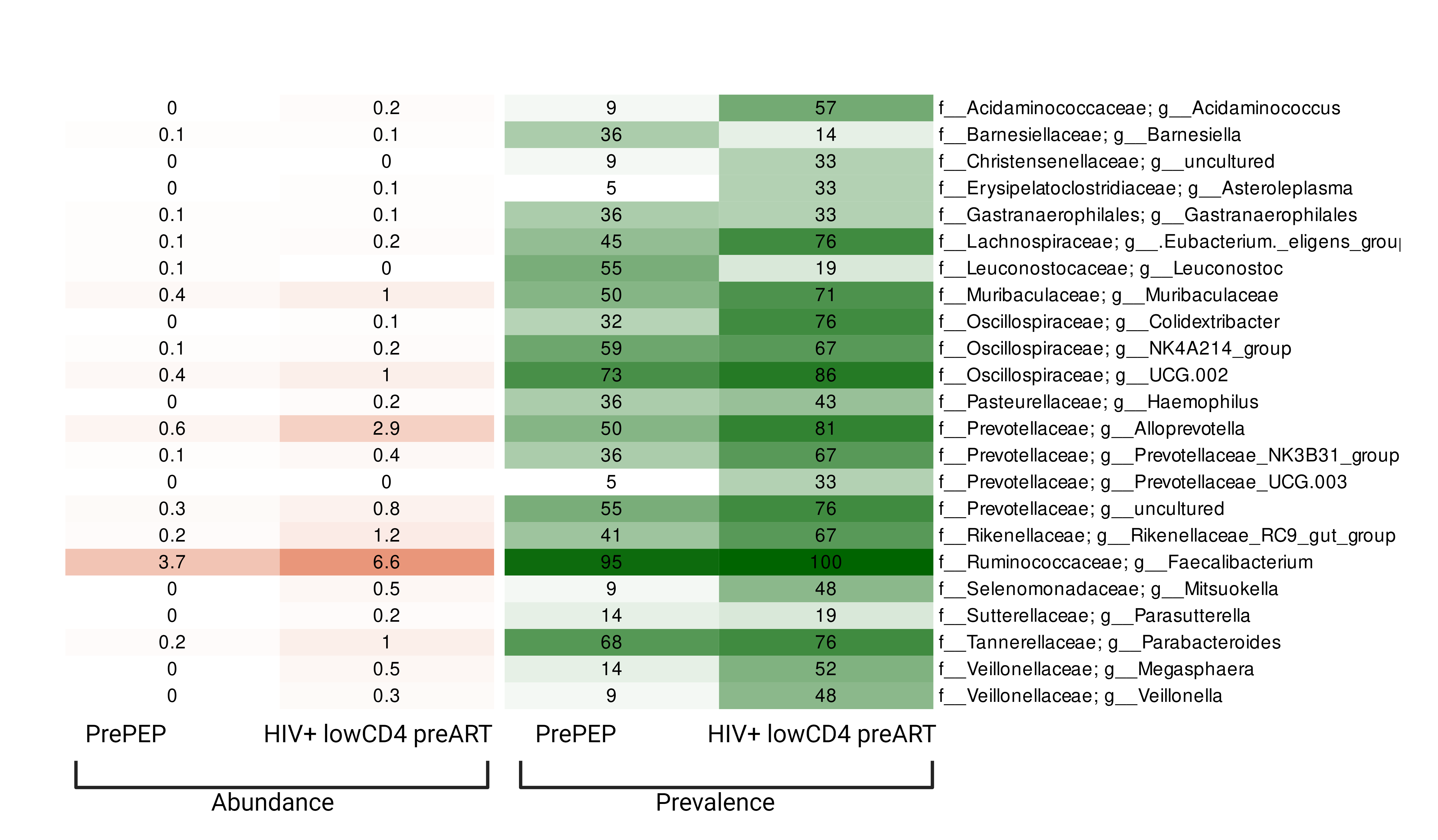

### **Figure S1: Relative abundance and prevalence of the differential bacterial taxa across groups.**

### ***Table S2: LFC of differential KO terms abundances between groups.***

| PrePEP vs HIV+ lowCD4 preART | | | | | | | |
| --- | --- | --- | --- | --- | --- | --- | --- |
| KO term     Description                            LFC       SE             P-value    Adjusted p | | | | | | | |
| K00992 | *murU; N-acetyl-alpha-D-muramate 1-phosphate uridylyltransferase* | | | 3.09 | 0.57 | 5.95 x10 ^-6^ | 2.96 x10 ^-2^ |
| K09477 | *citT; citrate: succinate antiporter* | | | 3.15 | 0.31 | 3.40 x10 ^-8^ | 1.73 x10 ^-4^ |
| K13040 | *ttrS; two-component system, LuxR family, sensor histidine kinase TtrS* | | 3.26 | | 0.44 | 1.15 x10 ^-7^ | 5.84 x10 ^-4^ |
| K12059 | *trbC; conjugal transfer pilus assembly protein TrbC* | -3.25 | | | 0.22 | 5.42 x10 ^-6^ | 2.69 x10 ^-2^ |
| HIV+ lowCD4 postART vs HIV+ highCD4 postART | | | | | | | |
| K17248 | *pglJ; N-acetyl galactosamine -N,N'-diacetylbacillosaminyl-diphospho-unde caprenol 4-alpha-N-acetyl galactosaminyl transferase* | 3.08 | | | 0.41 | 1.70 x10 ^-7^ | 8.69 x10 ^-4^ |
| K07100 | *K07100; putative phosphoribosyl transferase* | 3.06 | | | 0.29 | 4.53 x10 ^-7^ | 2.32 x10 ^-3^ |
| K13935 | *mdcH; malonate decarboxylase epsilon subunit [EC:2.3.1.39]* | -4.73 | | | 0.31 | 3.64 x10 ^-7^ | 1.86 x10 ^-3^ |
| PrePEP vs PostPEP | | | | | | | |
| K02840 | *waaB,rfaB;UDP-D-galactose:(glucosyl)LPS alpha-1,6-D-galactosyltransferase* | 2.70 | | | 0.14 | 1.08 x10 ^-6^ | 5.36 x10 ^-3^ |
| K13010 | *per, rfbE; perosamine synthetase* | -2.43 | | | 0.22 | 9.63 x10 ^-6^ | 4.78 x10 ^-2^ |

### ***Table S3: LFC of differential EC terms abundances between groups***

| **PrePEP vs HIV+ lowCD4 preART** | | | | | |
| --- | --- | --- | --- | --- | --- |
| **EC term          Description                                                           LFC             SE            p-value         Adjusted p** | | | | | |
| EC:1.1.1.320 | *Benzil reductase ((S)-benzoin forming)* | 2.79 | 0.25 | 2.97 x10 ^-5^ | 4.48 x10 ^-2^ |
| EC:3.4.24.84 | *Ste24 endopeptidase* | 2.58 | 0.38 | 4.17 x10 ^-7^ | 6.61 x10 ^-4^ |
| EC:5.3.3.10 | *5-carboxymethyl-2hydroxymuconate- isomerase* | -2.65 | 0.21 | 4.17 x10 ^-7^ | 6.61 x10 ^-4^ |
| EC:4.1.1.68 | *5-oxopent-3-ene-1,2,5-tricarboxylatedecarboxylase* | -2.77 | 0.23 | 2.39 x10 ^-6^ | 3.75 x10 ^-3^ |
| EC:1.2.1.39 | *Phenylacetaldehyde dehydrogenase* | -3.05 | 0.22 | 6.68 x10 ^-7^ | 1.06 x10 ^-3^ |
| **HIV+ lowCD4 postART vs HIV+ highCD4 postART** | | | | | |
| EC:2.4.1.291 | *4-alpha-N-acetylgalactosaminyltransferase* | 3.03 | 0.19 | 1.10 x10 ^-7^ | 1.79 x10 ^-4^ |
| EC:3.2.2.30 | *Aminodeoxyfutalosine nucleosidase* | 2.81 | 0.15 | 8.42 x10 ^-7^ | 1.36 x10 ^-3^ |
| **PrePEP vs PostPEP** | | | | | |
| EC:1.1.1.264 | *L-idonate 5-dehydrogenase* | 2.50 | 0.17 | 2.77 x10 ^-5^ | 4.38 x10 ^-2^ |
| EC:2.6.1.102 | *GDP-perosamine synthase* | -2.29 | 0.19 | 5.83 x10 ^-6^ | 9.23 x10 ^-3^ |
